# Sex Differences in Brain Structures and Functional Connectivity among Patients with Bipolar I Disorder

**DOI:** 10.1101/2021.03.29.21254597

**Authors:** Ming-Yang Li, Shih-Jen Tsai, Albert C. Yang

## Abstract

**Objectives:** Studies have demonstrated that sex differences may play a crucial role in the alternations of brain structures in individuals with bipolar disorder, but findings are not consistent. The current study identified sex differences in brain structure and function among a large sample of individuals with bipolar I disorder (BD-I).

**Methods:** Structural and functional magnetic resonance imaging datasets were acquired from 105 individuals with BD-I (36 men and 69 women) and 210 healthy adults (72 men and 138 women). A general linear regression model was used for voxel-wise analysis of grey matter (GM) and functional connectivity. Age, sex, diagnosis, and sex-by-diagnosis interaction were defined as predictors.

**Results:** In GM, the left caudate (*p* < .001), left thalamus (*p* < .001), right caudate (*p* = .003), right thalamus (*p* < .001), left anterior cingulate gyrus (*p* = .015), and left middle/posterior cingulate gyrus (*p* = .022) exhibited sex-by-diagnosis interaction. Furthermore, by using these six brain regions as seeds, we observed sex-by-diagnosis interaction in the alteration of functional connectivity between the left thalamus and right angular gyrus (*p* = .019).

**Conclusions:** Our data revealed a sex-by-diagnosis interaction associated with structure and function of the limbic system in individuals with BD-I. These findings may serve as reference for future studies on the pathophysiology of bipolar disorder.

## 1. Introduction

Bipolar disorder (BD) is characterized by episodes of depression and hypomania or mania and prolonged mood disturbances affecting cognition and other life functions. BDs are categorized according to the severity of manic symptoms into bipolar I disorder (BD-I) and bipolar II disorder (BD-II), with BD-I being characterized by manic episodes and BD-II being characterized by hypomanic episodes. The prevalence of BD is more than 1%, with BD-I predominating at a prevalence of 0.4%–0.6%.^1,2^ Relevant studies have revealed no sex differences in the prevalence, age of onset, or severity of BD symptoms.^3,4^ However, in clinical presentation, female individuals with BD have exhibited more frequent emotional state transitions than male individuals did, and female individuals often experienced depression.^3,5,6^ In addition, BD is often accompanied by sex-related comorbid symptoms, with anxiety disorders being frequent in women with BD^7^ and conduct and substance use disorders in men.^8,9^ Therefore, the sex differences related to the clinical statistics of BD identified in relevant literature may support the idea that brain structure and function in individuals with BD are influenced by sex factors.^10^

According to structural brain magnetic resonance imaging (MRI) studies, no sex differences in total brain volume of grey matter (GM) and white matter (WM) in individuals with BD have been observed.^4,10–13^ However, little is understood about the brain-related sex differences in individuals with BD; in particular, relevant literature is inconsistent regarding sex differences in the brain regions of individuals with BD.^10^ Evidence suggests that the volume of the right medial orbitofrontal cortex (OFC) is lower in men with BD than in women,^14^ and an increase in caudate volume was found in men with BD.^15^ Furthermore, men with BD exhibit low left anterior cingulate cortex (ACC) volumes, but no evidence of sex differences has been observed in other cingulate cortical subregions.^16^

Among functional MRI (fMRI) research, one study revealed that the activation state of the right ventrolateral prefrontal cortex was low in women with BD during fearful facial affect categorization.^17^ In addition, the activation state of the basal ganglia was high in women with BD during incentive decision-making.^18^ The study suggested that this occurred because women with BD have more frequent emotion-related transitions than men. However, only task-oriented fMRI studies have explored the sex differences in the activation state across brain regions in individuals with BD,^10,17,18^ and no resting-state fMRI study is available. Resting-state fMRI research on psychiatric individuals and healthy controls (HCs) is easily accessible and widely available and has been demonstrated to accurately indicate neural activity.^19,20^ Therefore, resting-state fMRI can be used to determine whether sex differences in nerve activity exist in individuals with BD at rest.

On the basis of the aforementioned literature, the current study established a systematic image analysis of sex-by-diagnosis interaction in brain structure and function by using a larger cohort of individuals with BD-I than relevant studies have used. Accordingly, the current study hypothesized that (1) sex differences exist in the brain structure of individuals with BD-I, and brain regions of interest (ROIs) can be identified, including OFC, ACC, and caudate;^14–16^ (2) the structural results of resting-state fMRIs can be used as brain ROIs to determine whether sex differences in resting-state neural activity exist.

## 2. Materials and Methods

### 2.1. Participants

The participants of the study were enrolled from the Taiwan Aging and Mental Illness Cohort (TAMI Cohort), which contains multimodality neuroimaging data from 587 HCs and 678 individuals with a major psychiatric disorder, and is jointly supported by Taipei Veterans General Hospital and National Yang Ming Chiao Tung University (NYCU).^21,22^ The brain images of all participants were scanned in the MRI room at NYCU, including structural MRI and resting-state fMRI. In addition, the TAMI Cohort contains demographic and clinical variable data.

The TAMI Cohort includes 112 individuals with BD-I (38 men and 74 women). Individuals were excluded for the following reasons: (1) they were left-handed (n = 2); (2) they scored lower than 26 on the Mini-Mental State Examination (MMSE) (n = 1);^23^ (3) they had symptoms of Parkinson’s disease (n = 1); and (4) demographic and clinical variable data were incomplete (n = 3). Finally, 105 individuals with BD-I were analyzed (36 men, mean age = 52.81 ± 15 years; 69 women, mean age = 45.93 ± 12.49 years). In total, 224 healthy adults corresponding to age and sex were extracted from the cohort at a ratio of 1:2. After screening, healthy adults were excluded for the following reasons: (1) they were left-handed (n = 5); (2) they scored lower than 26 on the MMSE (n = 2);^23^ and (3) they had a history of stroke or cerebral concussion and self-reported BD or depression (n = 7). Finally, a total of 210 HCs were analyzed (72 men, mean age = 47.68 ± 12.94 years; 138 women, mean age = 48.54 ± 14.63 years). The current study was approved by the Institutional Review Board of Taipei Veterans General Hospital and NYCU. All participants provided written informed consent. The protocol of the study is presented in Figure 1.

**Figure 1.**
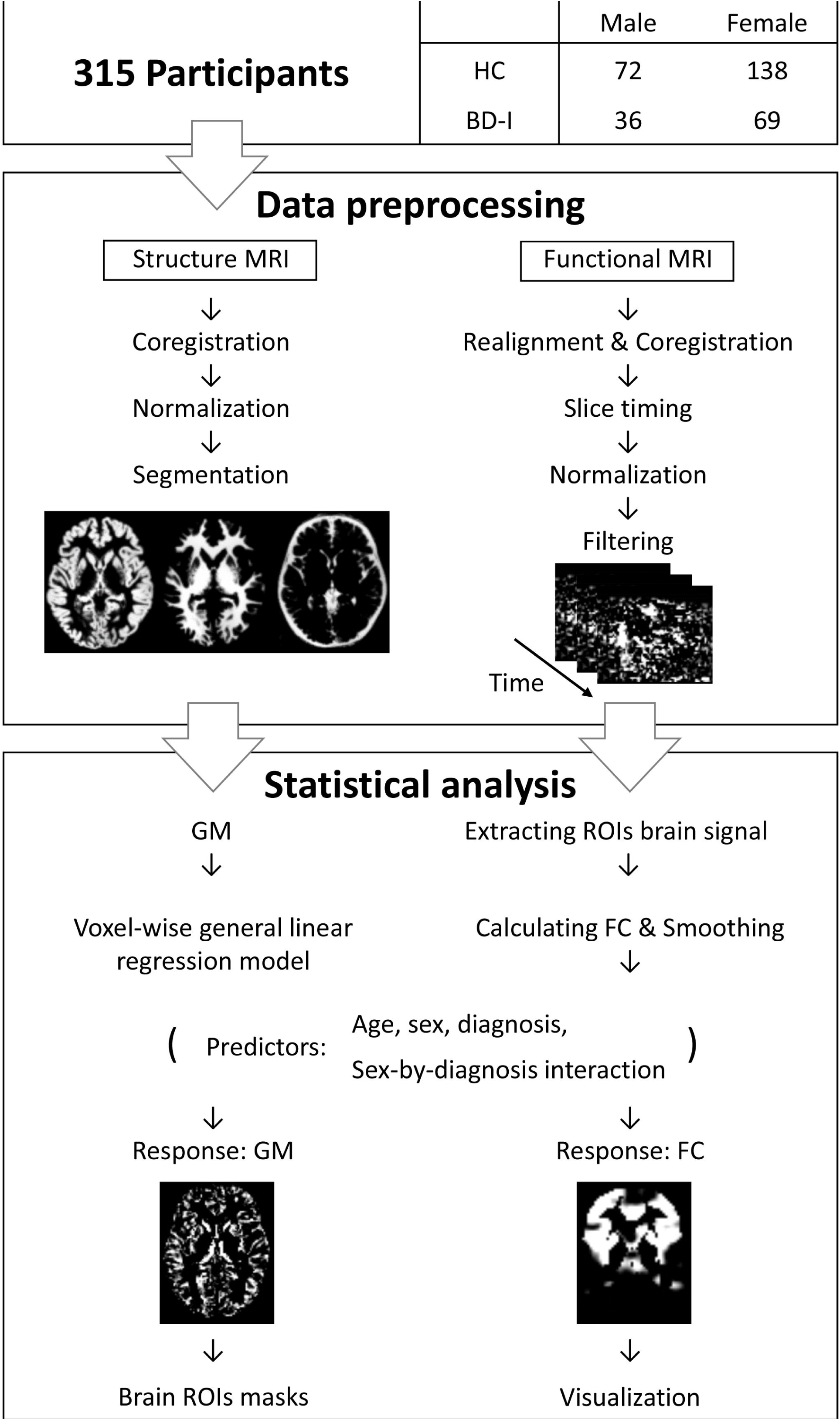
Flow chart. Abbreviations: HC, healthy controls; BD-I, bipolar I disorder; MRI, magnetic resonance imaging; GM, grey matter; ROI, region of interest; FC, functional connectivity

### 2.2. MRI Data Acquisition and Preprocessing

The MRI data of the participant were obtained from a 3.0-T Siemens MRI scanner with a 12-channel head coil. Participants were scanned for approximately 10 minutes with their eyes open and relaxed. All MRI data were visually reviewed by an experienced neuroradiologist to confirm that no morphologic abnormalities were present. Structural MRI and resting-state fMRI were processed using DPARSF_V4.3_170105^24^ and SPM12 under MATLAB 2019b ver. 9.7 (https://www.fil.ion.ucl.ac.uk/spm/software/spm12).

#### 2.2.1. Structural MRI

Whole-brain structural MRI data were collected using a three-dimensional (3D) magnetization-prepared rapid gradient echo sequence under the following imaging parameters: echo time (TE) = 3.5 ms; field of view = 256 ×256 mm^2^; voxel size = 1.0 × 1.0 × 1.0 mm^3^; flip angle 7°; and matrix size = 256 × 256. Preprocessing of structural MRI included head movement correlation, coregistering with resting-state fMRI, and segmentation and normalization to the Montreal Neurological Institute (MNI) space to generate GM, white matter (WM), and cerebral spinal fluid (CSF) images.

#### 2.2.2. Resting-State fMRI

Whole-brain resting-state fMRI data were collected using a T2*-weighted gradient-echo-planar imaging (EPI) sequence under the following imaging parameters: repetition time (TR) = 2500 ms, TE = 27 ms, field of view = 220 × 220 mm^2^, voxel size = 3.4 × 3.4 × 3.4 mm^3^, flip angle = 77°, matrix size = 64 × 64. In total, 200 EPI images were acquired along the anterior commissure–posterior commissure plane for each sequence. To eliminate the time difference between nerve activation and cerebral blood flow reaction, the first 5 of 200 time points were excluded. The number of slices was then set to 43, and the slice TR was set to 2.5 s to ensure accurate scanning time between slices. Next, tissue was realigned according to the participants’ structural MRI, and segmentation and normalization to the MNI space was performed according to the realigned tissue position. The blood flow signals of WM and CSF were used as covariates, and the component-based noise correction method and Derivative 12 head motion model were employed to eliminate irrelevant signals through nuisance regression. Finally, the band pass filter was set to 0.01–0.1 Hz for signal filtering.

### 2.3. Statistical Analysis

#### 2.3.1. Demographic and Clinical Characteristics

Group differences in demographic and clinical characteristics, including age, MMSE score, and rating on the Hamilton Depression Rating Scale (HAM-D) were evaluated through an independent *t* test by using SPSS ver. 24. The chi-squared test was also employed to compare the sex distributions within groups.

#### 2.3.2. Structural MRI

The GM images (voxel size = 2 × 2× 2 mm^3^) were imported into MATLAB by using the Neuroimaging Informatics Technology Initiative (NIfTI) toolbox.^25^ Normalization was performed using a template from the HCs of TAMI Cohort, and a general linear regression model (GLM) was then used. The probability of GM voxels was the dependent variable, and sex, age, diagnosis, and sex-by-diagnosis interaction were the predictors. The sex-by-diagnosis interaction statistics were visualized in xjView. Statistical significance was considered as follows: (1) voxel cluster = 30 voxels and (2) and voxel level *p* = .001. Brain regions with a cluster level *p* < .05 after adjustment for family-wise error rate were selected, and a brain ROI template was then created.

#### 2.3.3. Resting-State fMRI

Because the resolutions of structural MRI and resting-state fMRI differ, to extract the resting-state fMRI cerebral blood flow signal from the structural brain ROI template, the NIFTI toolbox was used to reslice the structural brain ROI template through nearest-neighbor interpolation to match the resolution of resting-state fMRI.

The resting-state fMRI images were imported into MATLAB by using the NIFTI toolbox (voxel size 3 × 3 × 3 mm^3^), and normalization was conducted using a template from the TAMI Cohort. The cerebral blood flow signals were extracted from the resliced structural brain ROI templates and subsequently averaged. The functional connectivity (FC) of cerebral blood flow signals between brain ROI and whole-brain GM voxels was calculated using Pearson correlation and Fisher transform. The calculated FC was used as the dependent variable in the GLM, and sex, age, diagnosis, and sex-by-diagnosis interaction were the predictors. The sex-by-diagnosis interaction statistics were visualized in xjView and BrainNet Viewer.^26^ Statistical significance was considered as follows: (1) voxel cluster = 30 voxels and (2) and voxel level *p* = .001. Brain regions with a cluster level *p* < .05 after adjustment for family-wise error rate were selected, and a brain map of resting-state FC was then created.

## 3. Results

### 3.1. Demographic and Clinical Characteristics

Table 1 presents the statistical results of the demographic and clinical characteristics of the individuals with BD-I and the HCs. Among individuals with BD-I, the mean age of male individuals was higher than that of female individuals (*p* = .01), and the mean severity of depression according to the HAM-D was higher in female individuals than in male individuals (*p* = .03). In addition, no significant sex differences were observed in the mean severity of mania (*p* = .48) and the percentage of individuals with psychotic characteristics (*p* = .2) according to the Young Mania Rating Scale. Among HCs, men had higher education levels than women did (*p* = .002), and no significant sex differences in mean age were observed (*p* = .67). Finally, a comparison of HCs with individuals with BD-I revealed that HCs had higher education levels and MMSE scores than individuals with BD-I (*p* = .002 and .001, respectively). No significant difference in mean age between the groups was identified (*p* = .98).

**Table 1.**
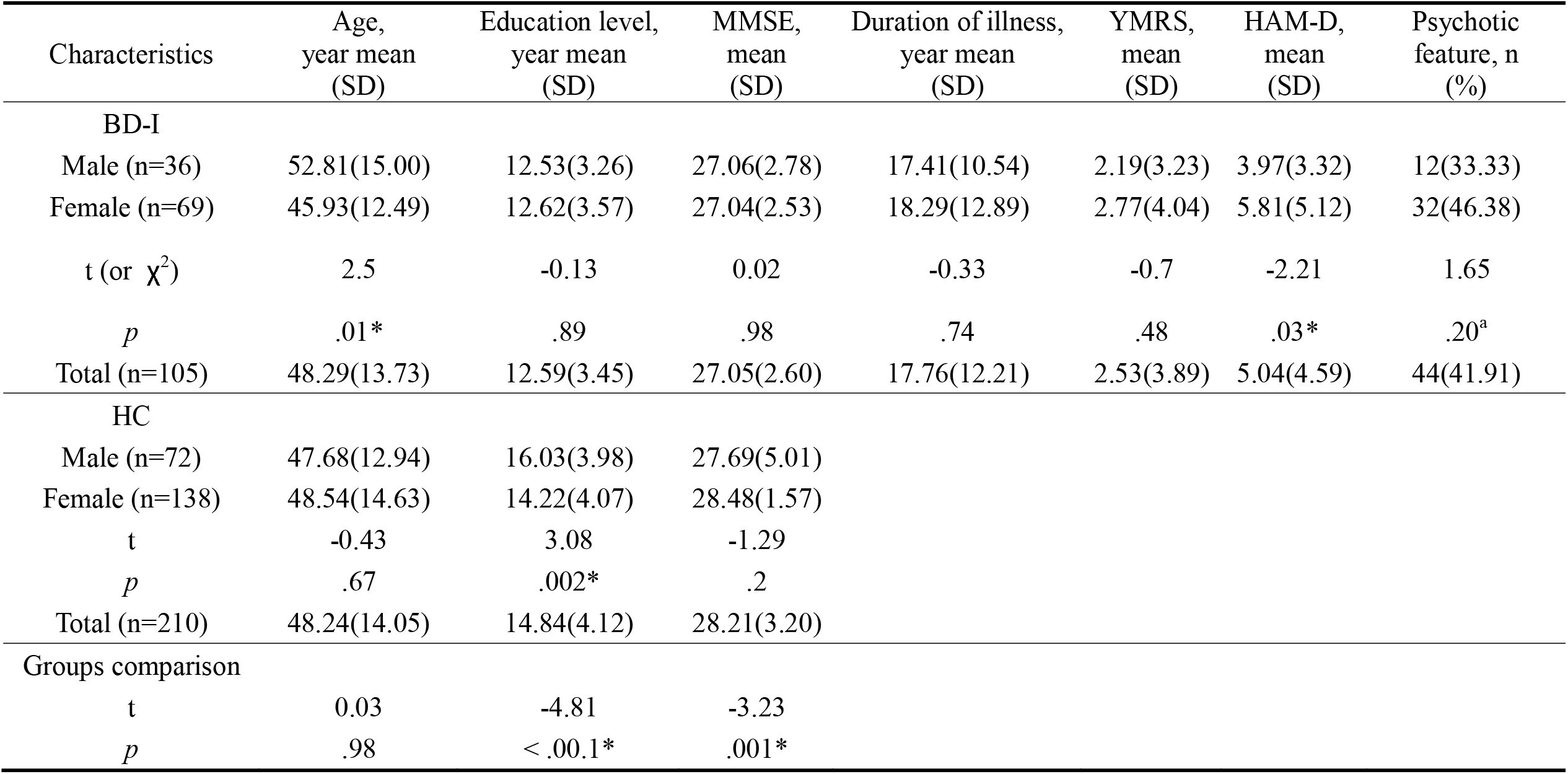
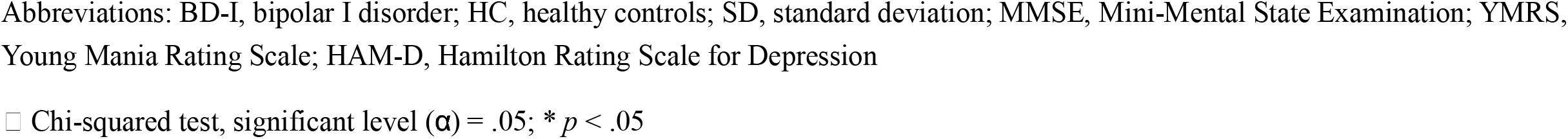
Demographic and clinical characteristics of bipolar I disorder and healthy controls.

| Characteristics | Age,<br>year mean<br>(SD) | Education level,<br>year mean<br>(SD) | MMSE,<br>mean<br>(SD) | Duration of illness,<br>year mean<br>(SD) | YMRS,<br>mean<br>(SD) | HAM-D,<br>mean<br>(SD) | Psychotic<br>feature, n<br>(%) |
| --- | --- | --- | --- | --- | --- | --- | --- |
| BD-I |  |  |  |  |  |  |  |
| Male (n=36) | 52.81(15.00) | 12.53(3.26) | 27.06(2.78) | 17.41(10.54) | 2.19(3.23) | 3.97(3.32) | 12(33.33) |
| Female (n=69) | 45.93(12.49) | 12.62(3.57) | 27.04(2.53) | 18.29(12.89) | 2.77(4.04) | 5.81(5.12) | 32(46.38) |
| t (or $\chi^2$) | 2.5 | -0.13 | 0.02 | -0.33 | -0.7 | -2.21 | 1.65 |
| p | .01* | .89 | .98 | .74 | .48 | .03* | .20<sup>a</sup> |
| Total (n=105) | 48.29(13.73) | 12.59(3.45) | 27.05(2.60) | 17.76(12.21) | 2.53(3.89) | 5.04(4.59) | 44(41.91) |
| HC |  |  |  |  |  |  |  |
| Male (n=72) | 47.68(12.94) | 16.03(3.98) | 27.69(5.01) |  |  |  |  |
| Female (n=138) | 48.54(14.63) | 14.22(4.07) | 28.48(1.57) |  |  |  |  |
| t | -0.43 | 3.08 | -1.29 |  |  |  |  |
| p | .67 | .002* | .2 |  |  |  |  |
| Total (n=210) | 48.24(14.05) | 14.84(4.12) | 28.21(3.20) |  |  |  |  |
| Groups comparison |  |  |  |  |  |  |  |
| t | 0.03 | -4.81 | -3.23 |  |  |  |  |
| p | .98 | < .00.1* | .001* |  |  |  |  |

Abbreviations: BD-I, bipolar I disorder; HC, healthy controls; SD, standard deviation; MMSE, Mini-Mental State Examination; YMRS, Young Mania Rating Scale; HAM-D, Hamilton Rating Scale for Depression
□ Chi-squared test, significant level ( $\alpha$ ) = .05; \* $p < .05$

### 3.2. Structural MRI

A GLM was used to determine the influence of the sex-by-diagnosis interaction on the GM of individuals with BD-I. Table 2 presents the brain regions with sex-by-diagnosis interaction as well as the coordinates of the brain regions with the most significant differences. The results drawn by the visualization software are presented in Figure 2, which displays a histogram of the brain region volume of HCs and the male and female individuals with BD-I.

**Table 2.** Sex-by-diagnosis interaction in voxel-wise statistics of grey matter among all participants.

| Brain regions | AAL | MNI coordinates (mm) |  |  | t | Cluster-level |  |
| --- | --- | --- | --- | --- | --- | --- | --- |
| | | x | y | z | | k | $p_{\text{FWE-corr}}$ |
| L caudate | 71 | -8 | 14 | -8 | 4.82 | 117 | < .001 |
| L thalamus | 77 | -6 | -10 | 8 | 4.95 | 130 | < .001 |
| L anterior cingulate gyrus | 31 | -4 | 32 | 12 | 4.79 | 31 | .022 |
| L middle/posterior cingulate gyrus | - | 0 | -30 | 28 | 4.41 | 34 | .015 |
| R caudate | 72 | 8 | 8 | -12 | 4.14 | 50 | .003 |
| R thalamus | 78 | 8 | -12 | 4 | 6.31 | 139 | < .001 |

Abbreviations: AAL, automated anatomical labelling; k, voxel numbers; $p_{\text{FWE-corr}}$ , $p$ after adjusting for multiple comparisons using a familywise error rate approach; L, left; R, right
Note: voxel size = 2x2x2 mm<sup>3</sup>

**Figure 2.**
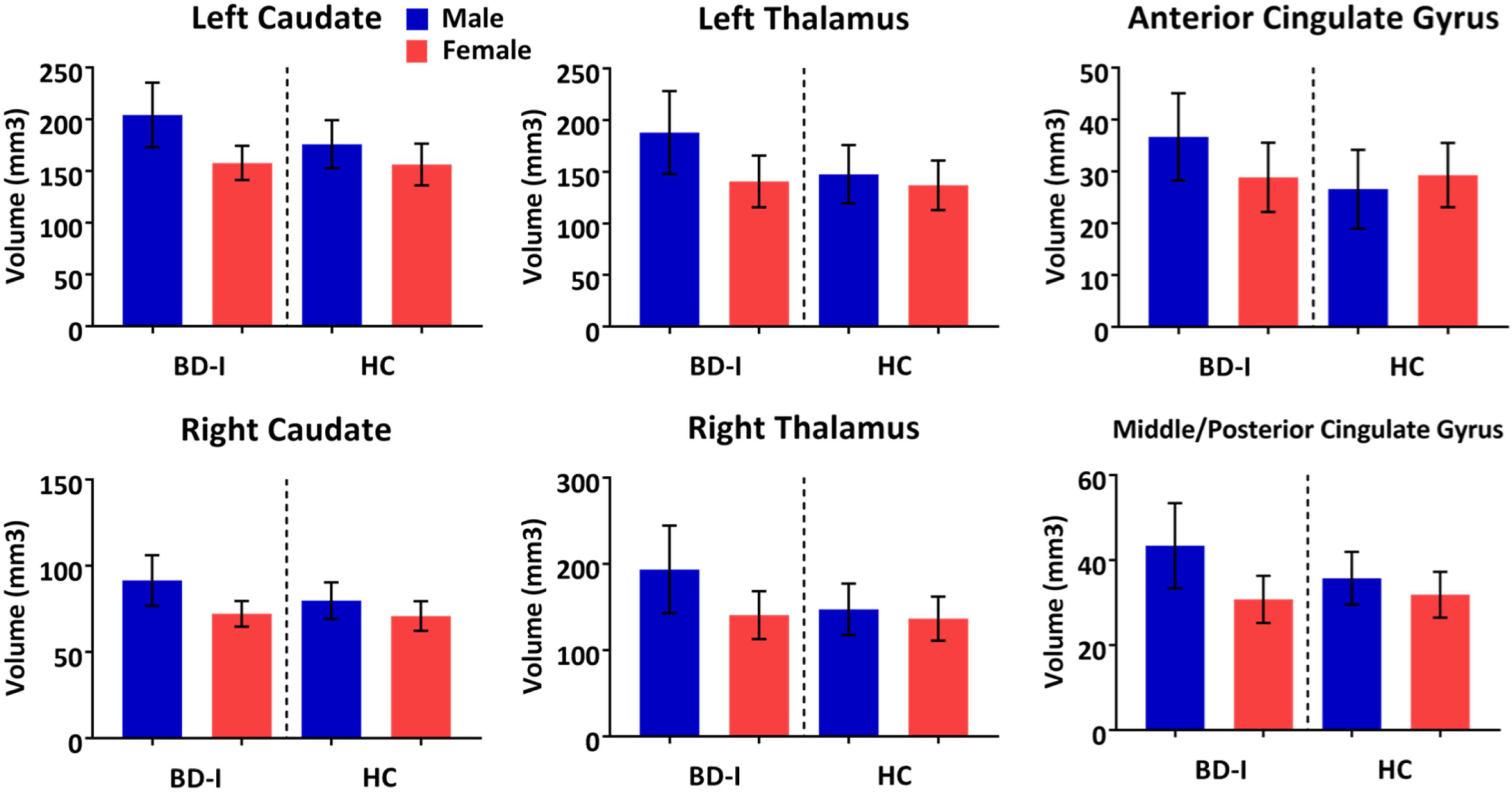
A histogram of brain region volume showing the sex-by-diagnosis interaction among all subjects. Abbreviations: HC, healthy controls; BD-I, bipolar I disorder

The brain regions depicted in Table 2 include the left caudate nucleus (*p* < .001), left thalamus (*p* < .001), left anterior cingulate gyrus (*p* = .022), left middle posterior cingulate gyrus (*p* = .015), right caudate (*p* = .003), and right thalamus (*p* < .001). Among them, the left middle posterior cingulate gyrus is located in the two brain regions of the automatic anatomical labeling (AAL) brain template; therefore, the AAL number is not indicated in Table 2. As presented in Figure 2, the caudate, thalamus, left anterior cingulate gyrus, and left middle posterior cingulate gyrus volumes in male individuals with BD-I are larger than those in female individuals and HCs.

### 3.3. Resting-State fMRI

We used the six brain ROIs that indicated sex-by-diagnosis interaction as the brain template to extract cerebral blood flow signals, and we averaged the extracted cerebral blood flow signals to calculate the FC with the whole-brain GM voxels. The GLM was then used to analyze the sex-by-diagnosis interaction. Table 3 presents the FC calculated using the left thalamus as seeds and the brain regions with sex-by-diagnosis interaction. The results drawn by the visualization software are presented in Figure 3, which displays a heat map of the significant differences in FC between male individuals with BD-I and female individuals and HCs.

**Table 3.** Sex-by-diagnosis interaction in voxel-wise statistics of functional connectivity using left thalamus seed among all participants.

| Brain regions | AAL | MNI coordinates<br>(mm) |  |  | t | Cluster-level |  |
| --- | --- | --- | --- | --- | --- | --- | --- |
| | | x | y | z | | k | $p_{\text{FWE-corr}}$ |
| R angular gyrus | 66 | 36 | -60 | 36 | 3.55 | 32 | 0.019 |
| R middle occipital gyrus | 52 | 39 | -69 | 33 | 3.20 | - | - |

Abbreviations: AAL, automated anatomical labelling; k, voxel numbers; $p_{\text{FWE-corr}}$ , $p$ after adjusting for multiple comparisons using a familywise error rate approach; L, left; R, right
Note: voxel size = 3x3x3 mm<sup>3</sup>

**Figure 3.**
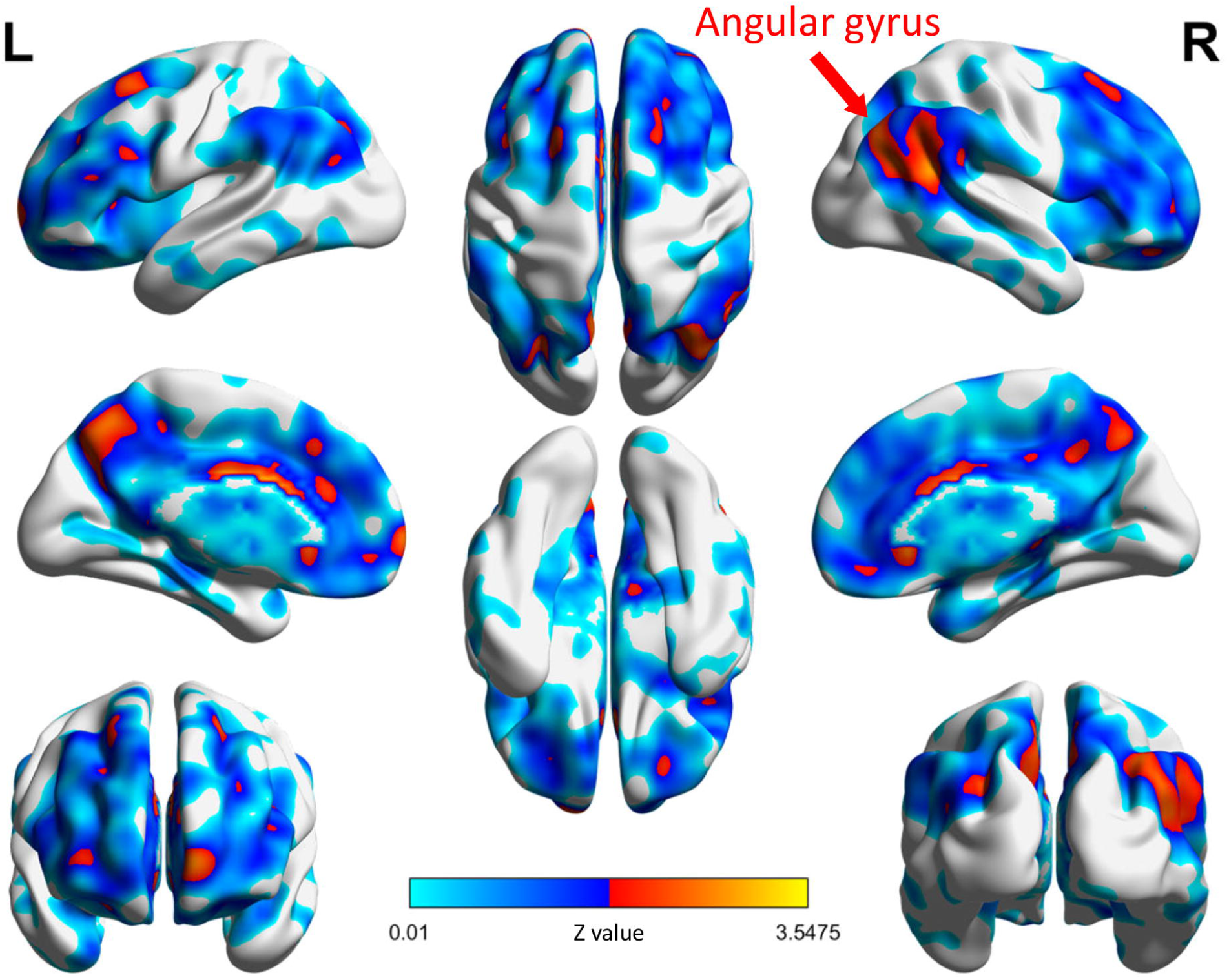
The visualized position of the brain region with the left thalamus as the seed, showing its functional connectivity according to the sex-by-diagnosis interaction.

Table 3 and Figure 3 indicate that the FC between the left thalamus and the right angular gyrus exhibited sex-by-diagnosis interaction, which was higher in male individuals with BD-I than in female individuals and HCs. No significant difference was observed in the FC between the other ROIs and the whole-brain GM voxels according to the GLM statistics.

## 4. Discussion

The current study used a substantial amounts of brain imaging data to identify potential sex differences in individuals with BD. The main findings were as follows: (1) the caudate, thalamus, left anterior cingulate gyrus, and left middle posterior cingulate gyrus volumes in male individuals with BD-I were larger than those in female individuals and HCs; and (2) the FC between the left thalamus and the right angular gyrus exhibited sex-by-diagnosis interaction.

### 4.1. Differences in the Clinical Characteristics of Individuals with BD-I

According to the statistical results, the severity of depression was higher in female individuals with BD-I than in male individuals. However, a relevant study suggested that no sex differences exist in terms of depression severity.^3^ The HAM-D classifies depression severity as a score indicating no depression (0–7), mild depression (8–16), moderate depression (17–23) and severe depression (≥24).^27^ Most individuals in the current study did not present with depression, but approximately 20% of female individuals (15 individuals) presented with mild or moderate depression. This statistical difference may be due to the higher scores of the female individuals. In addition, no significant sex differences were observed in mania severity and the percentage of individuals with psychiatric symptoms. This finding is consistent with relevant studies.^3,9^

### 4.2. Implication of Sex Differences in the Brain Structures of Individuals with BD-I

The statistical results of the GLM indicated that the caudate, thalamus, left anterior cingulate gyrus, and left middle posterior cingulate gyrus exhibited sex-by-diagnosis interaction. The caudate is a part of the basal ganglia and is associated with cognitive and emotional processing.^28^ Additionally, the thalamus and cingulate gyrus, which are both located in the limbic system, are also associated with emotional performance. The basal ganglia and limbic system are often used as key brain regions for BD research, and the brain ROIs discovered in the current study may also be key brain regions related to BD in terms of sex among individuals with BD-I.

First, the finding that the caudate exhibits sex-by-diagnosis interaction is consistent with relevant studies,^15^ and the increased caudate volume may contribute to the alleviation of impaired cognitive function.^29^ However, most studies have indicated that the caudate volume of individuals with BD is smaller than that of healthy people,^30–32^ but these results have suggested that individuals with BD have an abnormal basal ganglia structure. In addition, the structural abnormalities identified in the caudate may be related to the serotonin hypothesis.^33^ The availability of serotonin transporter in the caudate was observed to be lower in individuals with BD than in healthy people, indicating that individuals with BD do not transmit serotonin in normal amounts. Although this hypothesis has not been confirmed,^34^ it may serve as a possible direction for the further investigation of sex differences in individuals with BD-I.

Second, consistent with relevant studies, the left thalamus volume in individuals with BD is higher than that in healthy individuals.^35^ These results suggest that structural abnormalities of the left thalamus occur early in individuals with BD, possibly due to abnormal neuronal proliferation or pruning. In addition, male individuals with BD-I have been observed to have a higher amount of vesicular monoamine transporter protein (VMAT2) connected to the thalamus than female individuals and healthy individuals have. The concentration of VMAT2 may be associated with cognitive deficits in individuals with BD; in particular, reduced cognitive performance was observed in male individuals.^36^ In the current study, a sex-by-diagnosis interaction was observed in the thalamus, which may be explained by neuronal variation or changes in VMAT2 concentrations.

Moreover, the current study identified a sex-by-diagnosis interaction in the left anterior cingulate gyrus and left middle posterior cingulate gyrus. Relevant studies have identified increased anterior cingulate gyrus volumes in individuals with BD by using lithium salts.^37–39^ Lithium salts exhibit a nourishing effect on the nerves, protecting nerve cells, enabling them to grow, and restricting the persistent shrinking of the anterior cingulate gyrus.^39^ This is consistent with the clinical statistics reported in relevant studies that suggest that men use lithium for treatment more often than women.^6,40^ Such usage may have contributed to the larger cingulate gyrus volume in the male individuals in the current study.

Finally, the brain regions observed to exhibit sex-by-diagnosis interaction (i.e., the caudate, thalamus, left anterior cingulate gyrus, and left middle posterior cingulate gyrus) have been associated with cognitive function in individuals with BD.^29,36,41^ For example, caudate volume in individuals with BD was related to poor cognitive performance;^29^ poor verbal learning ability in male individuals with BD was related to the thalamus;^36^ and poor performance on the Wisconsin Card Sorting Test and Trail Making Test in individuals with BD was related to the anterior cingulate gyrus.^41^ These brain regions could be stimulated to improve cognitive function in individuals with BD according to sex.

### 4.3. Implications of Sex Differences in the FC of Individuals with BD-I

To our knowledge, only a few studies have used fMRI to explore sex differences in BD, most of which are task-oriented fMRI studies.^17,18^ The current study explored sex differences in BD-I through voxel-based analysis of resting-state fMRI and identified a sex-by-diagnosis interaction in FC between the left thalamus and the right angular gyrus. Individuals with schizophrenia exhibited increased FC in the thalamocortical network,^42^ including the thalamus and the angular gyrus. Currently, the thalamocortical network is mainly associated with rhythmic activities such as sleep.^43^ According to a relevant study, compared with healthy people, individuals with BD-I exhibited abnormal circadian rhythms.^44^ In addition, individuals with BD-I experience sleep problems, which were found to be related to manic episodes in female individuals.^45^ The possible sex differences in sleep among individuals with BD-I may involve the thalamocortex network, but evidence from follow-up studies is required to substantiate this explanation.

## 5. Limitations

This study had several limitations. First, relevant studies of structural brain images have also focused on WM. However, the current study did not analyze relevant WM changes related to sex-by-diagnosis interaction. Future studies may further explore the sex-by-diagnosis interaction between grey and white matter and its role in BD-I. Second, the current study is one of the few studies that used resting-state fMRI to identify sex differences in individuals with BD-I. According to the structural MRI results, the brain regions that exhibited sex-by-diagnosis interaction are associated with cognitive function. These brain regions could be used as brain ROIs for task-oriented MRI studies in future studies. In terms of methods, cerebral blood flow signals could be investigated using spectrum analysis to obtain more detailed information, which could contribute to a more comprehensive interpretation of results.

## 6. Conclusions

The current study established a systematic process for analyzing brain images to determine sex differences in the brain structure and FC in individuals with BD-I on the basis of the largest sample size obtained from relevant literature related to sex differences in individuals with BD-I. The caudate, thalamus, left anterior cingulate gyrus, and left middle posterior cingulate gyrus volumes were significantly higher in male individuals than in female individuals. Moreover, the FC between the left thalamus and the right angular gyrus exhibited a sex-by-diagnosis interaction, suggesting that alternations in brain function may be associated with sex differences in individuals with BD. Whether the results of the current study are related to the symptomological differences between the sex of individuals with BD-I remains unclear, and more evidence is required to confirm these findings. Future research should incorporate WM images and investigate resting-state fMRI through various signal analysis methods to confirm the findings of the current study.

## Data Availability

The data presented here are available from the corresponding author upon request.

## Acknowledgments

This work was supported by grants from the Ministry of Science and Technology of Taiwan [grant number 109-2628-B-010-011, 109-2321-B-010-006, and 109-2634-F-075-001 to Dr. Albert C. Yang and Dr. S. J. Tsai]. Dr. Albert C. Yang was also supported by Mt. Jade Young Scholarship Award from Ministry of Education, Taiwan, as well as Brain Research Center, National Yang Ming Chiao Tung University and the Ministry of Education (Aim for the Top University Plan), Taipei, Taiwan.

